# Risk factors for lung cancer in never-smokers: Multi-cohort study

**DOI:** 10.1101/2025.03.11.25323738

**Authors:** G. David Batty, Frederick K Ho, Steven Bell

## Abstract

**Background:** If lung cancer in never-smokers was a single disease entity, it would be the sixth most commonly occurring malignancy. Despite the population impact, its risk factors are poorly understood owing to a dearth of larger-scale, well-characterised studies.

**Methods:** We pooled individual-participant data from 18 prospective cohort studies comprising 91,588 never smokers (55,452 women) aged 16-102 years at study induction. Participants were linked to national death registries.

**Results:** A maximum of 17 years follow-up (mean 9.7) gave rise to 85 lung cancer deaths. Of the 19 potential determinants captured at baseline, only being older age (hazard ratio; 95% confidence interval per 10 year increase: 2.45; 2.11, 2.85), male (2.25; 1.46, 3.48), and having a high fruit and vegetable intake (2.29; 1.25, 4.17) were associated with elevated rates of lung cancer in this never-smoking group. No other substantial relationships were detected.

**Conclusions:** Despite the number and breadth of potential risk factors featured in this multi-cohort study, there was no clear suggestion of new determinants of lung cancer in never-smokers.

**Impact:** Our findings point to the need to explore the influence of risk factors additional to those included herein, particular in the field of genetics. Our unlikely finding for fruit and vegetable consumption warrants further testing.

## Introduction

Lung cancer is a leading cause of death worldwide and it is very well documented that cigarette smoking is its most powerful risk factor. Even with only 10% of lung cancer cases arising in people with no history of smoking,^1^ the absolute number of disease events is high, such that, if lung cancer in never-smokers was a single disease entity, it would be the sixth most commonly occurring malignancy.^2^ While this brings into sharp focus the need to identify the causes of lung cancer in never smokers, the process is methodologically challenging because large studies are required and electronic health records, the most obvious source of data, often lack information on smoking habit. The current evidence base implicates a history of non-cancer respiratory disease and exposure to second-hand smoke, asbestos, and radon as being among the few modifiable risk factors in the occurrence of lung cancer in people who never smoked.^1^ We therefore examined the role of an array of other determinants by pooling individual-participant data from a series of well-characterised, identical, field-based cohort studies.

## Methods

Described in detail elsewhere,^3^ we used individual-participant (raw) data from 18 cohort studies with identical methodology: the Health Survey for England (15 studies) and the Scottish Health Surveys (3 studies). Ethical approval for data collection was granted by the London Research Ethics Council and local research ethics committees, and study members consented to participate.

A total of 222,829 people participated in a home-based biomedical survey between 1994 and 2008 when aged 16-102 years; 193,873 consented to being linked to a national mortality registry (figure 1). Of these, 94,456 people reported that they had never smoked. The further omission of study members with a plasma or salivary cotinine >1.0 ng/mL to identify so called smoking deceivers;^4^ those reporting use of nicotine replacement therapy; and study members with a history of cancer, resulted in 91,588 (55,452 women) remaining individuals and this was our analytical sample.

**Figure 1.**
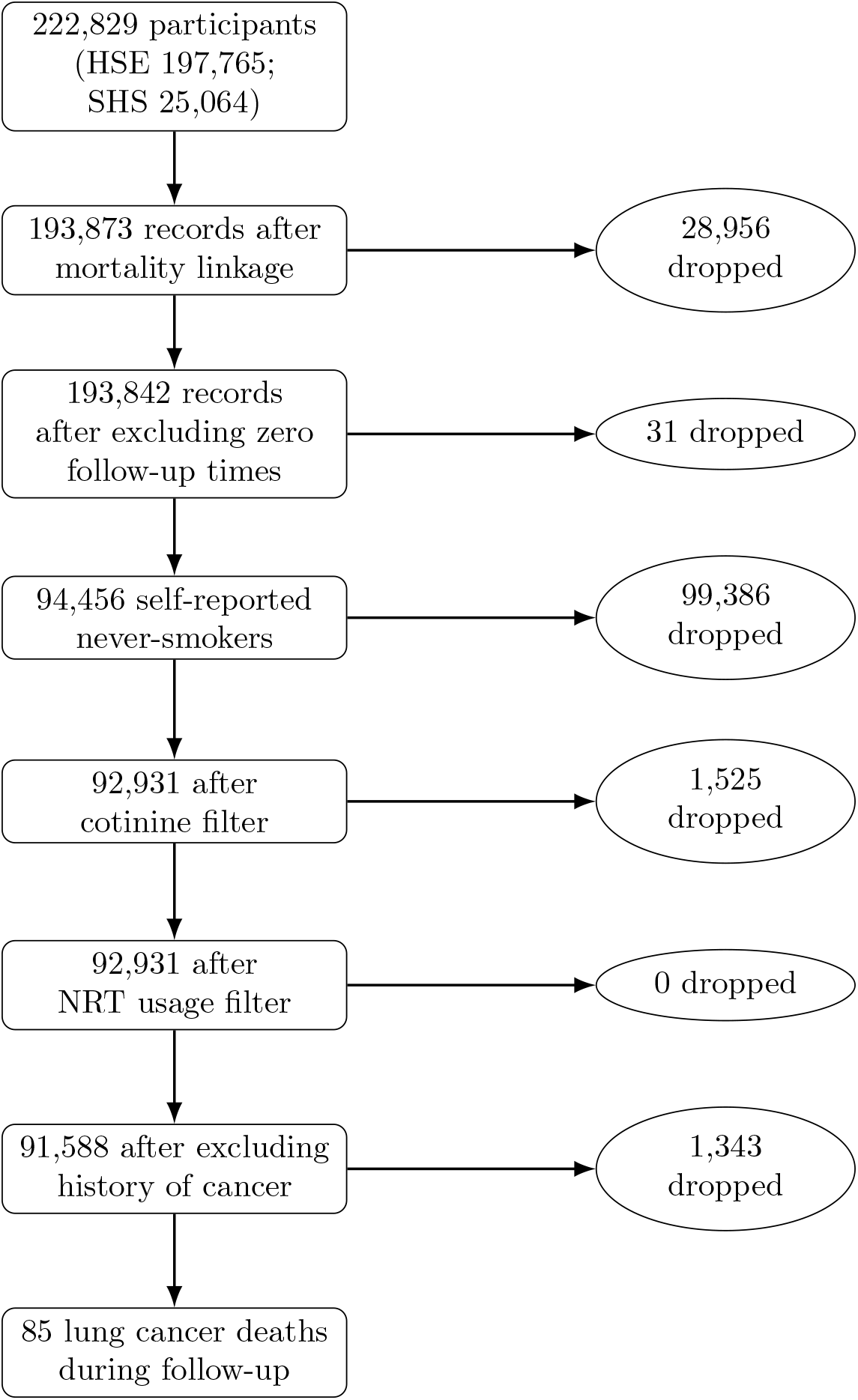
Flow of study members into the analytical sample: **Follow-up of study members in the Health Survey for England and the Scottish Health Surveys** HSE, Health Survey for England; SHS, Scottish Health Survey; NRT, nicotine replacement therapy

### Baseline data collection

At study baseline, health behaviours (dietary intake, including alcohol use, leisure-time physical activity), psychosocial characteristics (cohabitation status, educational attainment, psychological distress), physical health (self-rated general, respiratory medications, non-cancer respiratory illness), and demographic data (ethnicity, sex, age) were all self-reported using standard methods.^3^ During medical examination, waist and hip circumference, height and weight, C-reactive protein, and plasma or blood cotinine were also measured using standard protocols. To account for differences in age, sex, and height, forced expiratory volume in one second and forced vital capacity were standardised using the Global Lung Function Initiative 2022 equations^5^ and z-scores derived.

### Ascertainment of lung cancer mortality

Participants were linked to routinely-collected UK-wide mortality records from which cause of death was extracted. Ascertainment of lung cancer was based on any mention of the malignancy on the death certificate.^6^ A shared frailty Cox proportional hazards model accounted for unmeasured heterogeneity across studies by incorporating study-level random effects. Participants were censored according to the date of death from this malignancy or the end of follow-up (14 February 2011 in the Health Survey for England, 31 December 2009 in the Scottish Health Surveys), whichever came first. Analyses were conducted using Stata version 15.

## Results

A maximum of 17 years of mortality surveillance (mean 9.7) of 91,588 study members gave rise to 85 lung cancer deaths. Being older (hazard ratio; 95% confidence interval per 10 year increase: 2.45; 2.11, 2.85) and male (versus female: 2.25; 1.46, 3.48) was associated with an elevated risk of lung cancer (table 1). While there was also a suggestion of associations with some of the others study member characteristics – higher body mass index and alcohol consumption – these did not reach conventional levels of statistical significance. The only exception was higher intake of vegetable and fruit where there was a more than doubling in the rate of lung cancer mortality (2.29; 1.25, 4.17).

**Table 1.** Association between baseline characteristics and lung cancer mortality in life-long never smokers: Follow-up of study members in the Health Survey for England and the Scottish Health Surveys.

|  | Number of<br>people | Number<br>of deaths | Hazard ratio<br>(95% CI) | p-value |
| --- | --- | --- | --- | --- |
| **Socio-demographic factors** |  |  |  |  |
| Age (10-year change) | 91588 | 85 | 2.45 (2.11, 2.85) | <0.001 |
| Male vs female | 91588 | 85 | 2.25 (1.46, 3.48) | <0.001 |
| Non-white ethnicity vs white | 91530 | 85 | 0.79 (0.28, 2.23) | 0.65 |
| Low educational attainment vs higher | 91487 | 85 | 1.09 (0.69, 1.73) | 0.71 |
| Manual occupational social class vs non-manual | 84051 | 79 | 1.02 (0.65, 1.59) | 0.94 |
| **Health behaviours** |  |  |  |  |
| High fruit and vegetable consumption (≥5 servings/day) vs less | 91588 | 85 | 2.29 (1.25, 4.17) | <0.01 |
| Consumes alcoholic beverages vs abstinence | 90305 | 85 | 1.51 (0.77, 2.95) | 0.23 |
| Low leisure time physical activity vs higher | 56436 | 50 | 1.09 (0.61, 1.96) | 0.76 |
| Passive smoking (≥0.1 ng/mL blood/saliva cotinine) vs lower | 39384 | 32 | 0.86 (0.33, 2.25) | 0.75 |
| **Health history** |  |  |  |  |
| Poor self-reported general health vs higher | 91576 | 85 | 1.07 (0.46, 2.47) | 0.87 |
| High psychological distress (GHQ-12≥4) versus lower | 91588 | 85 | 1.26 (0.74, 2.15) | 0.39 |
| **Physiological factors** |  |  |  |  |
| Sex-standardised z-score for BMI | 82366 | 69 | 1.24 (0.97, 1.59) | 0.09 |
| Sex-standardised z-score for height | 86047 | 72 | 1.06 (0.83, 1.37) | 0.63 |
| Per doubling of C-reactive protein (mg/L) | 19007 | 24 | 1.04 (0.80, 1.34) | 0.78 |
| **Respiratory factors** |  |  |  |  |
| Any longstanding respiratory condition vs none | 91576 | 85 | 1.13 (0.52, 2.46) | 0.75 |
| Use of any respiratory medication vs none | 84838 | 80 | 1.19 (0.52, 2.74) | 0.68 |
| Global Lung Function Initiative z-score FEV1 | 25085 | 23 | 1.27 (0.91, 1.77) | 0.15 |
| Global Lung Function Initiative z-score FVC | 25085 | 23 | 1.26 (0.90, 1.76) | 0.18 |
| Global Lung Function Initiative z-score FEV1/FVC | 25085 | 23 | 1.14 (0.83, 1.57) | 0.42 |

Hazard ratios are age- and sex-adjusted. In selected survey years, no attempt was made to collect data on CRP, lung function, physical activity, and cotinine hence the lower sample size. GHQ-12, 12-item General Health Questionnaire; BMI, body mass index; FEV1, Forced Expiratory Volume in one second; FVC, Forced Vital Capacity

## Discussion

Our main finding was that, with the exception of risk factor associations for higher age and being male -- both well-replicated in this context^1^ -- there was little evidence of a clear association for the remaining 17 potential determinants of never-smoking lung cancer. Our finding that higher fruit and vegetable consumption appeared to confer elevated risk has been reported in other studies^7^ but is not a universal observation.^8^ Given the paucity of known risk factors for lung cancer in never smokers, we included as many potential determinants as possible from our dataset, however, rarely, some of these (e.g., psychological distress), were not hypothesis-driven.

In conclusion, despite the number and breadth of potential environmental risk factors described, there was no clear emergence of new determinants for never-smoking lung cancer. It may be that future research should consider the role of genetic indices.

## Data Availability

https://data-archive.ac.uk/

## Notes

Funding: GDB is supported by the US National Institute on Aging (1R56AG052519-01; 1R01AG052519-01A1) and SB by Cancer Research UK (A27657). There was no direct financial or material support for the work reported in the manuscript.

Conflicts of Interest. The authors declare no potential conflicts of interest.

### Competing Interest Statement

The authors have declared no competing interest.

### Funding Statement

GDB is supported by the US National Institute on Aging (1R56AG052519-01; 1R01AG052519-01A1) and SB by Cancer Research UK (A27657). There was no direct financial or material support for the work reported in the manuscript.

### Author Declarations

Ethical approval for data collection was granted by the London Research Ethics Council and local research ethics committees, and study members consented to participate.

